# Short Practical Regimen of Acupuncture for Melasma: A Prospective Cohort Study in a Tertiary Hospital in Thailand

**DOI:** 10.1101/2021.02.04.21251151

**Authors:** Thanan Supasiri, Nuntida Salakshna, Krit Pongpirul

**Author notes:** Co-Corresponding Authors Contact, Nuntida Salakshna, MD., Address: Samitivej Esthetics Institute, Samitivej Sukhumvit Hospital, 133 Sukhumvit 49, Klongtan Nua, Vadhana, Bangkok 10110 Thailand., Assist. Prof. Dr. Krit Pongpirul, MD, MPH, PhD., Address: Department of Preventive and Social Medicine, Faculty of Medicine, Chulalongkorn University, 1873 Rama IV Rd., Patumwan, Bangkok 10330, Thailand.

## Abstract

**Background:** Melasma is a common skin pigmentation disorder. Acupuncture shows benefits for patients with melasma, although no standardized method or an optimal number of sessions have been determined.

**Objective:** To comparatively assess clinical outcomes of melasma between five versus ten standard acupuncture sessions.

**Methods:** We performed a prospective observational cohort study in all melasma patients at the Acupuncture Clinic, King Chulalongkorn Memorial Hospital, Bangkok, Thailand. All patients were treated with acupuncture procedures twice a week and were evaluated after the fifth and the tenth sessions of acupuncture, with a one-week follow-up after the last session. Participants were divided into two treatment groups: Groups A and B received five and ten acupuncture sessions respectively. Melasma outcomes were assessed by using the Melanin Index (MI), Melasma Area and Severity Index (MASI), patient-reported improvement scores, and acupuncture-related adverse events.

**Results:** Of 113 participants, 67 received five sessions of acupuncture treatment while 39 received ten sessions. 7 participants did not finish treatment. At one week after five sessions of acupuncture in Group A, the mean MI decreased by 28.7 (95%CI -38.5 to -18.8, p<0.001), whereas the median MASI decreased by 3.4 (−6.9, -1.2), p<0.001) points. At one week after ten sessions of acupuncture in Group B, the mean MI decreased by 31.3 (95%CI -45.0 to -17.6, p<0.001), whereas the median MASI decreased by 5.4 (−9.9, -3.0), p<0.001) points. The first five sessions of acupuncture had a higher effect on the change in magnitude than the last five sessions, although there was no statistically significant difference. Twenty-nine participants reported minor side effects, mostly consisting of persistent pain and small hematoma. Group B had a risk ratio (RR) of having adverse events 1.8 times (95% CI 1.0-3.4, p-value = 0.05) compared to group A.

**Conclusion:** Short acupuncture regimen of 5 to 10 sessions in melasma seems to be effective, practical, and high patient satisfaction with minor side effects.

**Summary Box:** *What is already known about this subject?:*

- Acupuncture has effectively improved melasma condition at low cost with minimal side effects.
- Traditionally, one acupuncture course comprises of 10 sessions for a chronic condition but dose-response evidence of acupuncture for melasma has been lacking.
- Number of acupuncture sessions performed in various randomized controlled trial varied from 21 to 60 over 4-16 weeks.

*What are the new findings?:*

- Five sessions of acupuncture improved Melanin Index (MI) by 28.7 and Melasma Area and Severity Index (MASI) by 3.4 points.
- Ten sessions of acupuncture improved MI by 31.3 and MASI by 5.4 points, of which the improvement was mainly from the first five sessions.
- Short acupuncture regimen of 5 to 10 sessions in melasma seems to be effective, practical, and high patient satisfaction

*How might it impact on clinical practice in the foreseeable future?:*

- Dose-response study has not been common but could have clinical and financial implications.

## Introduction

Melasma is an acquired hyperpigmentation characterized by ill-defined light to dark brownish macules and patches, typically affect photo-exposed areas of the face.^1-3^ The most commonly implicated etiological factors are chronic ultraviolet exposure, female hormone stimulation, and genetic predisposition.^4-6^ Its appearance negative impacts the quality of life, emotional wellbeing, and even lead to psychiatric consequences.^7, 8^

In the last decade, there was a paradigm shift in the pathophysiology of melasma which is now considered to be more complex interplay between epidermal and dermal components including proliferation of dermal vessels and oxidative damage by reactive oxygen species.^4, 5^ Apart from conventional treatment such as hydroquinone, triple-combination cream (hydroquinone, tretinoin, and fluocinolone acetonide), and other bleaching agents, many new emerging therapies which targeting on vascular component such as tranexamic acid or vascular targeting laser and light treatment (such as pulsed dye laser and intense pulsed light) have shown benefits in lightening the melasma area.^9-12^

Acupuncture has shown benefits for patients with melasma. The effective rate of acupuncture for melasma has been reported around 87-96%.^14-16^ In Chinese medicine theory, acupuncture can promote blood and qi circulation, reducing blood stasis, rebalance and regulate various internal systems in the body to promote metabolism of epidermal cells, as well as regulate several hormones and antioxidants thus improving the melasma.^14, 17, 18^ Another RCT found that after acupuncture treatment for melasma, blood estrogen level decreased while progesterone increased, which corresponds to the association of increasing estrogen and decreasing progesterone level in melasma patients.^15^ Moreover, acupuncture for melasma has shown to be effective at low cost with minimal side effects which is beneficial for this chronic recurrent skin disease. However, it requires multiple frequent treatments, which can be inconvenient for the patients. Traditionally, one course of acupuncture treatment for a chronic disease consists of ten sessions,^21^ but the acupoints used and the optimal number of treatment sessions for acupuncture for melasma have been inconclusive. Therefore, this study aims to investigate and compare a short-fixed regimen of acupuncture between 5 and 10 sessions by body acupuncture with encircling and intralesional needling method.

## Methods

### Study design and population

This prospective observational cohort was conducted at the Acupuncture Clinic, King Chulalongkorn Memorial Hospital, Bangkok, Thailand, from September – December 2018. We recruited all melasma patients who came to the clinic for acupuncture treatment and were willing to participate in this observational study. We excluded participants who were pregnant, had severe underlying disease or psychosis, had a bleeding tendency, or had skin infections in the acupoints area. All participants were directed to maintain their usual behavior, such as using the same frequency and amount of sunscreen protection or continuing to not use sunscreen regularly.

### Acupuncture Procedure

All patients were treated with fixed acupoints: encircling the melasma area, Fengchi (GB20), Hegu (LI4), Xuehai (SP10), Jusanli (ST36), Sanyinjiao (SP6) and Taichong (LR3). All acupoints were performed bilaterally except those encircling the melasma area. The acupoints were carefully selected from the literature review of standard teaching materials in China^22^ and those frequently used in previous studies.^14, 20^ We used sterile disposable needles sized 0.25 × 40 mm. Acupuncture was done twice a week by well-trained acupuncturists, licensed with acupuncture Master’s degrees certified from China. “De Qi” sensation in every acupoint was mandatory, and needles were retained 30 minutes each time. Participants were evaluated every fifth session of acupuncture. Since this was an observational study, we used the usual protocol used in our acupuncture clinic. If the first five sessions of acupuncture did not yield satisfactory results and the patients were willing to continue the treatment, we continued the acupuncture treatment for five more sessions. Therefore, participants were divided into 2 groups: Group A received five acupuncture sessions and Group B received ten acupuncture sessions.

### Outcome Measurements

Participants were evaluated at the first visit, every five sessions of acupuncture, and one week after their last session.

The primary outcome was the change in Melanin Index (MI) measured by Mexameter^®^ MX 18 (Courage+Khazaka electronic GmbH, Köln, Germany) on melasma affected part of the face. MI was calculated as the average of a triplicate measurement in the area where the melasma was darkest.

The secondary outcomes consisted of the change in Melasma Area and Severity Index (MASI), patient-reported improvement scores and related adverse events from acupuncture. The MASI was developed by Kimbrough-Green et al^23^ and assesses three factors—area (A) of involvement, darkness (D) and homogeneity (H)—at four locations: the forehead (f), right malar region (rm), left malar region (lm) and chin (c). The MASI score can be calculated using the following formula:

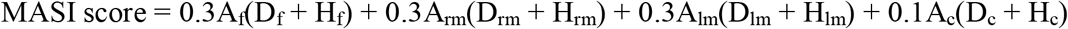

The area of involvement is rated from 0 to 6 while the darkness and homogeneity of pigmentation are rated from 0 to 4; thus, the MASI score ranges from 0 (none) to 48 (severe melasma).

Subjective improvement scores were measured by the six-point Likert Scale, from -1 to +4 (−1, worsening; 0, no improvement; +1, slight improvement; +2, moderate improvement; +3, marked improvement; +4, almost cured or completely cured). Adverse events due to acupuncture were recorded from interviewing participants and physical examinations by physicians each visit.

All participants were also photographed at every evaluation visit. MI, MASI and satisfactory scores were evaluated by the same evaluator who had been trained by a dermatologist.

### Statistical analysis

Baseline characteristics and data at each time-point were analyzed by descriptive statistics. Data after treatment compared to baseline in each group was analyzed by using the Student’s paired t-test or Wilcoxon signed-rank tests as appropriate. Linear regression was used to compare the results between Group A and Group B after adjusting for unequal baselines. Linear regression was also used to see if any baseline factors affected outcome. For Group B, mixed model with subject as a random effect was used to compare the result of the first five sessions (1^st^ to 5^th^) and the last five sessions (6^th^ to 10^th^). Pairwise comparison between the first and last five sessions was made by Scheffe’s Test. If the data did not meet the assumptions to use the mixed model or linear regression, the nonparametric test was used. The data were analyzed using the Stata/MP 15.1 (StataCorp LLC, College Station, TX, USA) software package.

### Trial Registration

The study was registered on August 19, 2018, before recruiting the first case, with the Thai Clinical Trials Registry, Identification Number TCTR20180824002.

### Ethics Approval

The study was approved by the Institutional Review Board (IRB), Faculty of Medicine, Chulalongkorn University (IRB No.544/2559). Each participant provided written informed consent.

## Results

One hundred and thirteen melasma patients came for acupuncture treatment during September – December 2018. Baseline characteristics are shown in Table 1. Acupuncturists finished the course of treatment after five sessions for 67 participants due to the significant improvement achieved. The other 39 participants continued until ten sessions. Seven participants did not complete at least five sessions of acupuncture and thus, were not evaluated.

**Table 1.** Characteristics of the Participants

|  | Group A:<br>Five sessions of acupuncture<br>treatment (n = 67) | Group B:<br>Ten sessions of acupuncture<br>treatment (n=39) |
| --- | --- | --- |
| Female [n (%)] | 64 (95.5) | 37 (94.9) |
| Age [mean (SD)] (years) | 44.3 (9.7) | 52.5 (9.7) |
| Duration of Melasma<br>[mean (SD)] (years) | 7.4 (6.8) | 8.3 (6.2) |
| Prior Acupuncture<br>Experience [n (%)] | 18 (26.9) | 13 (33.3) |
| Family History of Melasma<br>[n (%)] | 48 (71.6) | 22 (56.4) |

### Melanin Index (MI)

The average baseline MI for all participants at the darkest area was 298.9 (SD 79.7). Table 2 shows baseline MI and measurements at each time-point for Groups A and B.

**Table 2.** Mean Melanin Index (MI) at each evaluation visit

| Melanin Index | Group A (n=67)<br>Mean (SD) | p-value | Group B (n=39)<br>Mean (SD) | p-value |
| --- | --- | --- | --- | --- |
| Baseline | 287.4 (64.8) | Reference | 315.9 (98.5) | Reference |
| After the 5<sup>th</sup> session | 262.3 (62.5) | < 0.001* | 293.4 (90.6) | 0.001* |
| One week after 5<sup>th</sup> session | 258.7 (61.4) | < 0.001* |  |  |
| After the 10<sup>th</sup> session |  |  | 285.6 (94.7) | < 0.001* |
| One week after 10<sup>th</sup> session |  |  | 284.6 (89.2) | < 0.001* |

\*p-value calculated using Student's paired t-test

In Group A, after the 5^th^ session of acupuncture, MI decreased by 25.0 (95%CI -34.0 to - 16.1, p<0.001): from 287.4 (SD 64.8) to 262.3 (SD 62.5). At one week after completing five sessions, the mean MI decreased by 28.7 (95%CI -38.5 to -18.8, p<0.001) to 258.7 (SD 61.4) from baseline. In Group B, after the 5^th^ session of acupuncture, MI decreased by 22.5 (95%CI - 35.5 to -9.5, p=0.001): from 315.9 (SD 98.5) to 293.4 (SD 90.6). After the 10^th^ session of acupuncture, MI decreased by 30.3 (95%CI -42.4 to -18.2, p<0.001) to 285.6 (SD 94.7) from baseline. At one week after completing ten sessions of acupuncture, the mean MI decreased by 31.3 (95%CI -45.0 to -17.6, p<0.001) to 284.6 (SD 89.2) from baseline.

Comparing the efficacy of the first five sessions (1^st^ to 5^th^) to the last five sessions (6^th^ to 10^th^) in Group B, the MI decreased by 22.5 (95%CI -35.5 to -9.5) and 7.8 (95%CI -20.5 to 4.8) respectively. However, there was no statistically significant difference between these two periods (p-value from Scheffe’s Test = 0.44).

From the linear regression analysis of the post-fifth-session treatment outcomes, apart from the different baseline MI between groups, other factors including the duration of melasma, prior acupuncture experience and family history of melasma did not affect the treatment outcome (see appendix 1).

Between the two groups at one-week follow up post-treatment, after adjusting for the unequal baseline, there was no significant difference between the two groups (p=0.67) (see appendix 2).

### Melasma Area and Severity Index (MASI)

The baseline median MASI scores for overall participants was 6 (2.6, 11.4) scores. Group B had a higher baseline MASI score than Group A, as shown in Table 3 with details of the MASI at each time point.

**Table 3:** Median Melasma Area and Severity Index (MASI) Changes from Baseline

| MASI | Group A<br>(n=67)<br>Median (IQR) | p-value | Group B<br>(n=39)<br>Median (IQR) | p-value |
| --- | --- | --- | --- | --- |
| Baseline | 5.2 (2.4, 9.0) | Reference | 8.1 (4.8, 14.4) | Reference |
| After the 5<sup>th</sup> session | 1.5 (1.2, 3.0) | < 0.001 | 4.8 (1.8, 8.0) | < 0.001 |
| One week after 5<sup>th</sup> session | 1.2 (0.6, 2.1) | < 0.001 |  |  |
| After the 10<sup>th</sup> session |  |  | 1.8 (1.2, 4.6) | < 0.001 |
| One week after 10<sup>th</sup> session |  |  | 1.4 (1.1, 4.4) | < 0.001 |

\*p-value from Wilcoxon signed-rank test

In Group A, after completing five sessions of acupuncture, the median change of MASI score was -2.9 (−5.3, -0.9) scores. Moreover, at one week post-fifth session, the median change of MASI score compared to baseline was -3.4 (−6.9, -1.2) scores. In Group B, after completing five sessions of acupuncture, the median change of MASI score was -3.3 (−6.0, -0.7) scores. After completing ten sessions of acupuncture, the median change of MASI score compared to baseline was -5.1 (−8.9, -2.5) scores. The median change of MASI score between the 6^th^ to 10^th^ session was -1.2 (−3.0, -0.6) scores. At one week after completing ten sessions of acupuncture, the median change of MASI score was -5.4 (−9.9, -3.0) scores.

Due to the non-normal distribution of the MASI scores, the Wilcoxon rank sum test was used to compare the change of MASI score between the two groups (p-value=0.08).

### Patient-Reported Melasma-Improvement

The frequency distribution of patient-reported improvement scores at each time point is shown in Table 4. After full treatment in both groups, 95.5% of Group A and 97.4% of Group B felt markedly improved or cured.

**Table 4.** Patient-Reported Improvement Scores

| *Satisfactory score* | *Group A (n=67)* |  | *Group B (n=39)* |  |
| --- | --- | --- | --- | --- |
|  | *n* | *%* | *n* | *%* |
| *After the 5<sup>th</sup> session* |  |  |  |  |
| *Worsening* | - | - | - | - |
| *No improvement* | - | - | - | - |
| *Slight improvement* | - | - | 2 | 5.7 |
| *Moderate improvement* | 3 | 4.5 | 4 | 11.4 |
| *Marked improvement* | 23 | 34.3 | 9 | 25.7 |
| *Almost or Completely Cured* | 41 | 61.2 | 20 | 57.1 |
| *After the 10<sup>th</sup> session* |  |  |  |  |
| *Worsening* |  |  | - | - |
| *No improvement* |  |  | - | - |
| *Slight improvement* |  |  | - | - |
| *Moderate improvement* |  |  | 1 | 2.6 |
| *Marked improvement* |  |  | 7 | 18.0 |
| *Almost or Completely Cured* |  |  | 31 | 79.5 |

### Related Adverse Events from Acupuncture

There were no major adverse events. Of 106 participants, 29 (27.4%) participants reported minor side effects (Table 5), 14 (20.9%) participants were from group A and 15 (38.5%) participants were from group B. Five patients reported two symptoms: two had hematoma and pain, two had pain and dizziness, and the last had headache and sweating. The other 24 participants had one discomfort symptom. Group B had risk ratio (RR) of having adverse event 1.8 times (95% CI 1.0-3.4, p-value = 0.05) compared to group A. All participants were willing to continue the treatment despite the adverse events.

**Table 5.** Adverse Events from Acupuncture reported by 29 out of 106 Participants

|  | Group A<br>(n = 67) |  | Group B<br>(n = 39) |  |
| --- | --- | --- | --- | --- |
|  | Events | % | Events | % |
| *Pain* | 7 | 10.4 | 8 | 20.5 |
| *Small hematoma* | 7 | 10.4 | 3 | 7.7 |
| *Dizziness* | 1 | 1.5 | 2 | 5.1 |
| *Headache* | 1 | 1.5 | 1 | 2.6 |
| *Sweat* | 1 | 1.5 | 1 | 2.6 |
| *Fatigue* | - | - | 1 | 2.6 |
| *Hot flush at right arm area* | 1 | 1.5 | - | - |

## Discussion

Acupuncture has shown clinical benefit for melasma through MI, MASI and patient-reported improvement. Both groups’ MI and MASI scores showed significant improvement after five sessions of acupuncture treatment. More than 95% of all participants subjectively reported marked improvement, which corroborates other previous studies’ high effective rate of acupuncture treatment for melasma.^14, 15^

Since melasma is a chronic recurrent skin disease, the short practical regimen of acupuncture has many positive clinical and financial implications as in other chronic diseases. White A et al^24^ mentioned adequate dosage for acupuncture, and some meta-analysis studies proposed the adequacy criteria of at least six acupuncture sessions for low back pain^25^ and chronic knee pain. ^26^ Sangtin et al applied a survival analysis technique in their retrospective observational data and found that 50% of stroke patients had significant improvement after eight sessions of acupuncture and reached peak improvement by sixteen sessions.^27^ For melasma, previous systematic review of 8 RCTs found the number of acupuncture sessions varied from 21 to 60 over 4–16 weeks (2–7 times weekly).^20^ In real practice, coming to the hospital multiple times per week for a long period of time is not practical both for the patients and hospital staffs. In our study, MI and MASI scores significantly decreased after 5-10 sessions with more than 95% of patients reported markedly improved or cured. Moreover, the first five sessions seem to have more magnitude of improvement than 6^th^ to 10^th^ sessions, although without statistically significant difference. Thus, given our setting, our protocol of 5-10 sessions is more practical than other studies’ protocol of more than 20 sessions,^20^ while it is still effective with high patient’s satisfaction.

We realized that from Chinese medicine theory, one of the important basic theories is syndrome differentiation and treatment according to each syndrome. However, we found that our short fixed regimen of acupoints also yielded effective result for the majority of patients, also making the treatment easier to train and perform. The treatment was likely effective because our selected acupoint included encircling and intralesional melasma area, resulting in a local increase of blood flow.^19^ This correlates with evidence from an RCT’s finding that acupuncture can increase skin blood perfusion.^19^ Moreover, a recent systematic review of melasma treatment which targeting on vascular components by tranexamic acid and laser/light therapies showed promising evidence.^3, 9, 11^ Apart form the blood circulation and vascular component which are the recent target treatment in melasma, a latest evidence revealed that melasma is a photoaging disorder^5, 6^ which related to oxidative damage by reactive oxygen species.^4^ Antioxidant properties of acupuncture in melasma also correlates with promising evidence for the use of antioxidants to deal with melasma in modern medicine.^14, 28^ Apart from the local effects, acupoints in the rest of the body acupoint—Fengchi (GB20), Hegu (LI4), Xuehai (SP10), Jusanli (ST36), Sanyinjiao (SP6) and Taichong (LR3)—have already been established to improve commonly seen syndromes of melasma patients: liver-qi stagnation, spleen-deficiency and kidney-yin/yang-deficiency.^18, 29^

In our study, the minor yet frequent adverse events of acupuncture were pain and hematoma in the facial area. Therefore, when conducting acupuncture on the facial area, clinicians should apply pressure after taking off the needles for longer period of time. Higher sessions of treatment seemed to have higher rate of adverse events.

Although this study is an observational study which collected data in a pragmatic manner, important factors including duration of melasma, prior acupuncture experience and family history of melasma had no effect on the clinical outcome of the treatment according to the regression result. However, this study had a relatively short follow up time. Further research should be conducted to see how long the effects last.

## Conclusions

Short acupuncture regimen of 5 to 10 sessions in melasma seems to be effective, more practical, and high patient satisfaction with minor side effects.

## Data Availability

Data is available upon reasonable request.

## Acknowledgement

The authors would like to thank Dr. Thanapoom Rattananupong, Assoc. Prof. Somrat Lertmaharit and Dr.Yuda Chongpison for statistical analysis advice.

## Contributorship

TS, NS and KP conceived of the idea, designed the study, collected and analyzed the data, drafted and approved the manuscript.

## References

1. Kauh YC, Zachian TF. Melasma. Advances in experimental medicine and biology. 1999;455:491–9.

2. Miot LD, Miot HA, Silva MG, Marques ME. [Physiopathology of melasma]. Anais brasileiros de dermatologia. 2009;84(6):623–35.

3. Masub N, Nguyen JK, Austin E, Jagdeo J. The Vascular Component of Melasma: A Systematic Review of Laboratory, Diagnostic, and Therapeutic Evidence. Dermatol Surg. 2020;46(12):1642–50.

4. Sarkar R, Bansal A, Ailawadi P. Future therapies in melasma: What lies ahead? Indian J Dermatol Venereol Leprol. 2020;86(1):8–17.

5. Kwon SH, Na JI, Choi JY, Park KC. Melasma: Updates and perspectives. Exp Dermatol. 2019;28(6):704–8.

6. Passeron T, Picardo M. Melasma, a photoaging disorder. Pigment Cell Melanoma Res. 2018;31(4):461–5.

7. Deshpande SS, Khatu SS, Pardeshi GS, Gokhale NR. Cross-sectional study of psychiatric morbidity in patients with melasma. Indian journal of psychiatry. 2018;60(3):324–8.

8. Handel AC, Miot LD, Miot HA. Melasma: a clinical and epidemiological review. Anais brasileiros de dermatologia. 2014;89(5):771–82.

9. Perper M, Eber AE, Fayne R, Verne SH, Magno RJ, Cervantes J, et al. Tranexamic Acid in the Treatment of Melasma: A Review of the Literature. Am J Clin Dermatol. 2017;18(3):373–81.

10. Del Rosario E, Florez-Pollack S, Zapata L, Jr., Hernandez K, Tovar-Garza A, Rodrigues M, et al. Randomized, placebo-controlled, double-blind study of oral tranexamic acid in the treatment of moderate-to-severe melasma. J Am Acad Dermatol. 2018;78(2):363–9.

11. Hassan AM, Elfar NN, Rizk OM, Eissa NY. Pulsed dye laser versus intense pulsed light in melasma: a split-face comparative study. J Dermatolog Treat. 2018;29(7):725–32.

12. Kong SH, Suh HS, Choi YS. Treatment of Melasma with Pulsed-Dye Laser and 1,064-nm Q-Switched Nd:YAG Laser: A Split-Face Study. Ann Dermatol. 2018;30(1):1–7.

13. Jutley GS, Rajaratnam R, Halpern J, Salim A, Emmett C. Systematic review of randomized controlled trials on interventions for melasma: an abridged Cochrane review. Journal of the American Academy of Dermatology. 2014;70(2):369–73.

14. Rerksuppaphol L, Charoenpong T, Rerksuppaphol S. Randomized clinical trial of facial acupuncture with or without body acupuncture for treatment of melasma. Complement Ther Clin Pract. 2016;22:1–7.

15. Wu X, Xiang Y. The Effects of Acupuncture Combined with Auricular Acupressure in the Treatment of Chloasma. Evidence-based complementary and alternative medicine : eCAM. 2018;2018:6438458.

16. Xun J. Clinical observation in 30 cases of chloasma treated by auricular point pressing and pricking. Journal of traditional Chinese medicine = Chung i tsa chih ying wen pan. 2003;23(3):207.

17. Chen W. Fifty cases of chloasma treated by acupuncture plus intensive pulse light irradiation. Journal of traditional Chinese medicine = Chung i tsa chih ying wen pan. 2007;27(4):265–7.

18. Feng XJ, Fu JY, Liu F. Clinical observation on the combined use of acupuncture and herbal medicine for treatment of chloasma. Journal of traditional Chinese medicine = Chung i tsa chih ying wen pan. 2010;30(1):15–7.

19. Kim K-W, Hong S, Kim HS, Kim T, Ahn J, Song H-S, et al. Physiological impact of nanoporous acupuncture needles: Laser Doppler perfusion imaging in healthy volunteers. PLOS ONE. 2019;14(12):e0226304.

20. Chai Q, Fei Y, Cao H, Wang C, Tian J, Liu J. Acupuncture for melasma in women: a systematic review of randomised controlled trials. Acupuncture in medicine : journal of the British Medical Acupuncture Society. 2015;33(4):254–61.

21. 王启. ▪ 灸治▪学. 10 ed. Beijing: 中国中医▪出版社; 2009.

22. Wang Q. Zhen Jiu Zhi Liao Xue (Book in Chinese). 10th ed. Beijing: Zhong Guo Zhong Yi Yao Chu Ban She.

23. Kimbrough-Green CK, Griffiths CE, Finkel LJ, Hamilton TA, Bulengo-Ransby SM, Ellis CN, et al. Topical retinoic acid (tretinoin) for melasma in black patients. A vehicle-controlled clinical trial. Archives of dermatology. 1994;130(6):727–33.

24. White A, Cummings M, Barlas P, Cardini F, Filshie J, Foster NE, et al. Defining an adequate dose of acupuncture using a neurophysiological approach – a narrative review of the literature. Acupuncture in Medicine. 2008;26(2):111.

25. Manheimer E, White A, Berman B, Forys K, Ernst E. Meta-analysis: acupuncture for low back pain. Annals of internal medicine. 2005;142(8):651–63.

26. White A, Foster NE, Cummings M, Barlas P. Acupuncture treatment for chronic knee pain: a systematic review. Rheumatology (Oxford, England). 2007;46(3):384–90.

27. Sangtin S, Supasiri T, Tangsathitporn R, Pongpirul K. Application of survival analysis techniques to determine the optimal number of acupuncture therapy sessions for stroke patients. Acupuncture in medicine : journal of the British Medical Acupuncture Society. 2020;38(3):194–200.

28. Babbush KM, Babbush RA, Khachemoune A. The Therapeutic Use of Antioxidants for Melasma. J Drugs Dermatol. 2020;19(8):788–92.

29. Hu JS. Acupuncture treatment of chloasma. Journal of traditional Chinese medicine = Chung i tsa chih ying wen pan. 2008;28(4):305–7.

