## Appendix 1, 2 for "Short Practical Regimen of Acupuncture for Melasma: A Prospective Cohort Study in a Tertiary Hospital in Thailand"

**Appendix 1: Linear regression analysis of the post-fifth-session treatment MI outcomes and the baseline characteristics of the participants**

| **Predictor** | **Effect Estimate** | **95% Confidence Interval** | **p-value** |
| --- | --- | --- | --- |
| **Intercept** | 31.13 | [-0.45, 62.72] | 0.05 |
| **Baseline MI** | -0.17 | [-0.26, -0.08] | <0.001 |
| **Duration of melasma (per year)** | 0.43 | [-0.67, 1.52] | 0.42 |
| **Prior acupuncture experience** |  |  |  |
| Never | Ref |  |  |
| Have previous acupuncture experience | -1.08 | [-16.61, 14.46] | 0.89 |
| **Family history of melasma** |  |  |  |
| No family history of melasma | Ref |  |  |
| 1^st^ degree relative have melasma | -11.61 | [-26.88, 3.66] | 0.13 |

**Appendix 2: Linear regression analysis of the one-week follow up post-treatment MI outcome comparing between the 2 groups after adjusting for unequal baseline MI**

| **Predictor** | **Effect Estimate** | **95% Confidence Interval** | **p-value** |
| --- | --- | --- | --- |
| **Intercept** | 31.21 | [2.93, 59.49] | 0.03 |
| **Baseline MI** | -0.21 | [-0.30, -0.12] | <0.001 |
| **Group** |  |  |  |
| A | Ref |  |  |
| B | 3.31 | [-12.00, 18.62] | 0.67 |
